# Effect of background therapy with Non-steroidal anti-inflammatory drugs (NSAIDs) and other anti-inflammatory agents on COVID-19 outcomes

**DOI:** 10.1101/2024.12.07.24318645

**Authors:** Lucio Miele, San Chu, William Hillegass, Claudine Jurkovitz, William Beasley, David Chen, A Jerrod Anzalone, Daniel Fort, John Kirwan, Brian Melancon, Sally Hodder, Ronald Horswell, the National COVID Cohort Collaborative Consortium

## Abstract

**Background:** Inflammation plays a complex, incompletely understood role in the pathogenesis of acute COVID-19 and Post-Acute Sequelae of SARS-CoV-2 infection (PASC or “Long COVID”). Systemic acute inflammation resulting in cytokine storm, hypercoagulability and endothelial damage is thought to be a central mechanism for severe morbidity and mortality in acute COVID-19. Anti-inflammatory medications taken routinely for chronic conditions prior to contracting COVID-19 (“background medications”) may modulate acute COVID-19 outcomes.

**Methods:** Using data from the National COVID Cohort Collaborative (N3C) enclave, we estimated effects of six classes of background medications on acute COVID outcomes. Medication classes included aspirin, celecoxib, other NSAIDS, steroids, immune suppressants, and antidepressants. Acute COVID outcomes included probability of hospital admission, inpatient mortality, and mortality among diagnosed COVID patients. Each medication class was compared to benzodiazepines (excluding midazolam) which served as a comparator/control. Only adult COVID patients with pre-existing osteoarthritis and without any diagnosed autoimmune disease were included in the analyses. Random effects logistic regression models were used to adjust for covariates and data contributing organization. Medication effects also were estimated for COVID-negative cases.

**Results:** Non-aspirin NSAIDS were associated with lower mortality among diagnosed COVID-19 patients: adjusted Odds Ratio (aOR)=0.32 (p=.032) for celecoxib; aOR=0.51 (p<.001) for NSAIDS other than aspirin and celecoxib. For inpatient mortality: aOR=0.34 (p=.060) for celecoxib and aOR=0.74 (p=.200) for other non-aspirin NSAIDS. Similar effects were observed for COVID-negative cases, including for inpatient mortality: aOR=0.21 (p<.001) for celecoxib and aOR=0.34 (p<.001) for other non-aspirin NSAIDS. Secondary analyses examined alternative explanations for results.

**Discussion:** Protective effects were observed for non-aspirin NSAIDS, especially celecoxib. However, those estimated effects implicitly assume the medication classes did not differ on the probability a true COVID-19 case was diagnosed. The similarity of COVID-positive and COVID-negative results suggest possible missing covariates. However, such similarity plausibly could stem from a medication having both “direct” and “indirect” effects on COVID outcomes. Adjudicating among the alternative interpretations would require data beyond those available. However, the effects observed for non-aspirin NSAIDS, while possibly biased, rationalize further investigation using study designs constructed to overcome the limitations of existing datasets.

## 1. Introdtiuction

Inflammation plays a complex and incompletely understood role in the pathogenesis of acute COVID-19 [1–4] and Post-Acute Sequelae of SARS-CoV-2 infection (PASC or “Long COVID”) [5]. Systemic acute inflammation resulting in cytokine storm, hypercoagulability and endothelial damage is thought to be a central mechanism for severe morbidity and mortality in acute COVID-19 [1–4].

Several classes of anti-inflammatory agents have been proposed as COVID-19 treatments. A meta-analysis of 7 randomized controlled trials (RCT) showed that corticosteroids (dexamethasone, hydrocortisone or methylprednisolone) decrease mortality when used to treat critically ill COVID-19 patients [6].

Non-steroidal anti-inflammatory drugs (NSAIDs) have been considered potentially unsafe in COVID-19 [7] due to their cardiovascular and renal side effects and the potential to decrease anti-viral responses [7], though very little information is available on their safety in COVID-19 patients [8]. At least two selective serotonin re-uptake inhibitors (SSRIs), fluvoxamine and sertraline, were suggested to be effective as adjunctive therapy for COVID-19 based on their anti-inflammatory effects [9].

The anti-inflammatory agents of interest here were used not as therapy for acute events but rather as “background medications” (i.e., medications taken routinely for chronic conditions). Here we examine the possible effects on COVID outcomes of several background medication groups, all of which have anti-inflammatory activity.

Using the National COVID Collaborative Cohort (N3C) database [10], Reese et. al. [11] focused on analyzing the effect of NSAIDs (including celecoxib, diclofenac, droxicam, etodolac, ketorolac, ibuprofen, indomethacin, lornoxicam, meloxicam, naproxen, piroxicam, tenoxicam) on patients with hospital admissions for COVID-19, finding a statistically significant relationship between NSAIDs use prior to admission and inpatient mortality (odds ratio = 0.51), but noted the potential vulnerability of that estimate to missing covariates.

The research question that our analyses seek to address is: Is there an association between background chronic treatment with anti-inflammatory medications and COVID-19 outcomes? To address that question, we conducted analyses using N3C data, as described below, for osteoarthritis patients who did not have concomitant autoimmune disorders, as these patients are most likely to be taking prescription anti-inflammatory agents and are not subject to the possible confounding effects of autoimmune disorders. Within the osteoarthritis group, we also considered patients taking immune-suppressant medications (e.g., transplant recipients) and patients taking SSRIs or systemic, oral steroids, based on the literature on these agents in COVID-19.

The estimation of background medication treatment effects is challenging for several reasons, leading to estimated treatment effects whose interpretation is ambiguous. Much of this paper is devoted to development of strategies to help clarify interpretation of estimated medication effects.

The analyses and results described in this paper were generated by using the National Institutes of Health’s National COVID Cohort Collaborative (N3C) database. N3C is a centralized, secure harmonized electronic health record database containing U.S. nationwide COVID-19-related data. As of April 2023, the N3C database included data from 74 data partner (DP) healthcare organizations. Each DP has contributed data for two types of individuals: (1) Individuals with at least one positive COVID-19 test result or provider diagnosis (here called “COVID-positive” patients), and (2) individuals who were tested for COVID-19 one or more times but with no positive test results on record (called “COVID-negative” patients).

## 2. Methods

### 2.1. Eligibility

Each DP contributed clinical data for all its identified COVID-19 positive cases. Each DP also contributed a sample of its COVID-19 negative cases, with that sample generated by matching to COVID-19 positive cases on age, gender, race, and ethnicity using a positive-to-negative matching ratio of 1:2. The N3C data were first made available for research in August 2020 and have been subsequently updated weekly. The data sent to N3C by each DP include demographics, symptoms, lab test results, diagnoses, procedures, medications, medical conditions, physical measurements. Of the 74 DPs contributing data to N3C, data contributions from 70 DPs were of potential use. As of February 12, 2022 (the date of data extraction for this project) those 70 DPs had contributed data for 12,093,403 patients. Data from 70 of the 74 DPs were used. The four excluded DPs’ data could not be used, as those DPs’ data lacked explicit dates of events (e.g., outcomes events) that the analysis required.

The diagram in Fig 1 summarizes how an ultimate group of 485,779 osteoarthritis patients was selected from those 12,093,403 patients. To avoid the possible confounding effects of vaccination on COVID outcomes, only data covering events during the time span 01/01/2020 through 03/31/2021 (i.e., before widespread vaccination implementation) were used. To assure the cohort included only patients with well-documented clinical histories, only adult patients with outpatient visits spanning 30+ days and prescription information spanning 30+ days were considered potentially eligible.

**Figure 1:**
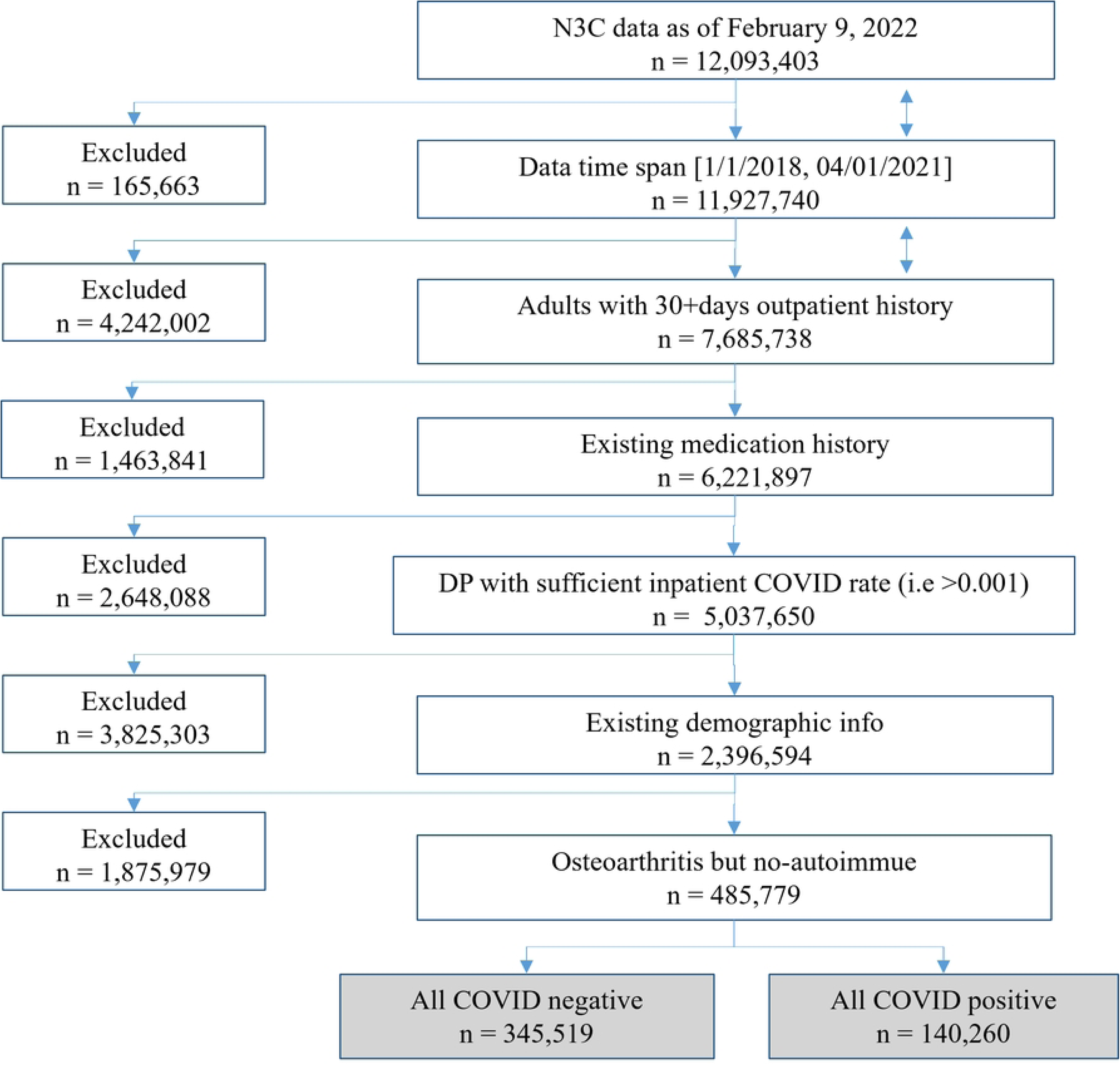
Selection of patients meeting eligibility criteria.

Some of the N3C DPs do not have inpatient facilities, and their data contributions include little (inpatient COVID-19 rate ≤ 0.001) or no inpatient data. Therefore, data from four (of 74) DP organizations were excluded, because the outcomes measures used in our analyses cannot be defined without use of inpatient information. Exclusions for age <35 and for missing demographic information left 2,396,594 individuals eligible before clinical exclusions. To achieve a more nearly clinically homogenous cohort of patients, but a cohort with substantial use of anti-inflammatory agents, eligibility was further restricted to patients with diagnosed osteoarthritis but without autoimmune disease. Specifically excluded for autoimmune disease were those diagnosed with rheumatoid arthritis, lupus, juvenile rheumatoid arthritis, inflammatory bowel disease, as well as certain other less prevalent autoimmune disorders. Further, regardless of diagnosis codes present, any patients using medications strongly associated with autoimmune disease treatment were excluded. The medications whose use led to exclusion included DMARDs, Methotrexate, cytokine-targeted biologics, non-anti-inflammatory biologics, and anti-allergy biologics. The above inclusion/exclusion criteria led to a final group of 485,779 osteoarthritis patients, of which 140,260 were “COVID-positive” and 345,519 were “COVID-negative.”

### 2.2. Background medication (treatment) groups

The anti-inflammatory medications considered represent six distinct medication groupings groups, as shown in Table 1. In addition, a seventh medication group (benzodiazepine sedatives) was defined to serve as a control group. Benzodiazepines were selected because to our knowledge they lack clinically meaningful anti-inflammatory or anti-viral effects. As depicted by the solid arrows in Fig 2, the analyses described here compare each of the six anti-inflammatory medication groups to the control group. Inclusion in a medication group required evidence (in the form of a prescription date) that the medication was used both within the 30 days prior to COVID-19 diagnosis and also more than 30 days prior to the COVID-19 diagnosis.

**Table 1:** Background Medication Groups.

| Medication group | Descriptions |
| --- | --- |
| **Non-specific NSAIDs** | Non-steroidal anti-inflammatory drugs (excludes celecoxib and aspirin) |
| **Celecoxib** | Celecoxib was considered individually due to its selectivity for COX-2 as opposed to other prescription NSAIDs |
| **Aspirin** | Aspirin was considered individually as it is often prescribed for its anti-platelet activity to patients at significant risk for cardiovascular or cerebrovascular disease. |
| **Immune suppressants** | Small-molecule, non-steroid immune suppressants often prescribed to prevent rejection in transplant recipients were included due to their potential suppressive effects on anti-viral immunity. |
| **Steroids** | Steroids were included due to their potential suppressive effects on anti-viral immunity. |
| **Antidepressants** | Antidepressants (SSRIs and tricyclic medications) have anti-inflammatory properties and can be prescribed for osteoarthritic pain. |
| **Benzodiazepine sedatives**<br>**(excluding midazolam)** | **(This group serves as a comparator /control group.)**<br>As a small-molecule control class, we selected benzodiazepine sedatives. These are prescribed as anxiolytics or hypnotics. To our knowledge, this class of medications has no known clinically significant anti-inflammatory or anti-viral effects. |

**Figure 2:**
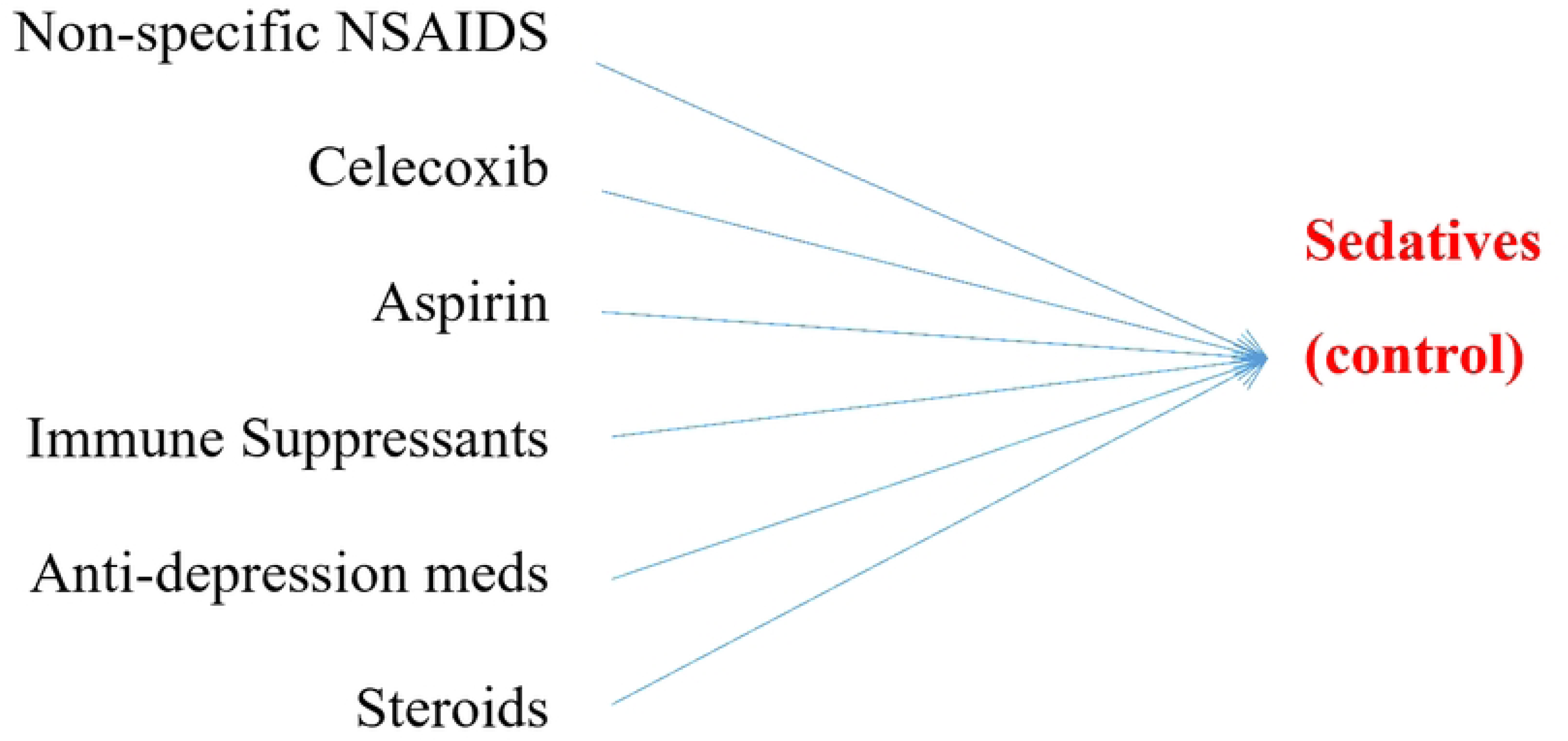
Pairwise Comparisons used in the Primary Analyses.

### 2.3. Primary analyses

Each of the six pairings with the control medication group shown in Fig 2 led to six distinct statistical models, which estimated six comparison measures which for convenience we refer to as M1, M2, M3, M4, M5, and M6. After adjustment for demographics and known comorbidities, three of the measures compare outcomes among COVID-positive patients, and three compare outcomes among COVID-negative patients. Specifically, the COVID-positive measures are:

**M1** = the adjusted odds ratio (aOR) comparing **90-day death rates**, defined as death or discharge to hospice within the 90 days following the first date on which the subject was known to be COVID-positive.

**M2** = the adjusted odds ratio comparing **90-day admission rates**, defined as a hospital admission within the 90 days following the first date on which the subject was known to be COVID-positive.

**M3** = the adjusted odds ratio comparing **death rate among hospitalized patients**, defined as death or discharge to hospice within the 90 days following a hospital admission, with that admission required to have occurred within +/- seven days of the subject’s first known COVID-positive date.

The three corresponding measures for COVID-negative patients are:

**M4** = the adjusted odds ratio comparing **90-day death rates**, defined as death or discharge to hospice within the 90 days following an encounter date randomly selected from the subject’s encounters.

**M5** = the adjusted odds ratio comparing **90-day admission rates**, defined as a hospital admission within the 90 days following an encounter date randomly selected from the subject’s encounters.

**M6** = the adjusted odds ratio comparing **death among hospitalized patients,** defined as death or discharge to hospice within 90 days of a hospital admission, with the hospital admission randomly selected from among the subject’s hospitalizations.

Because many patients use multiple medications, each of the above six comparison measures was estimated using the exclusion logic depicted in Fig 3. For example, when comparing aspirin users to benzodiazepine users (the control medication group), those using both of those medications were excluded. Also excluded were those using any of the other Table 1 medications.

**Figure 3:**
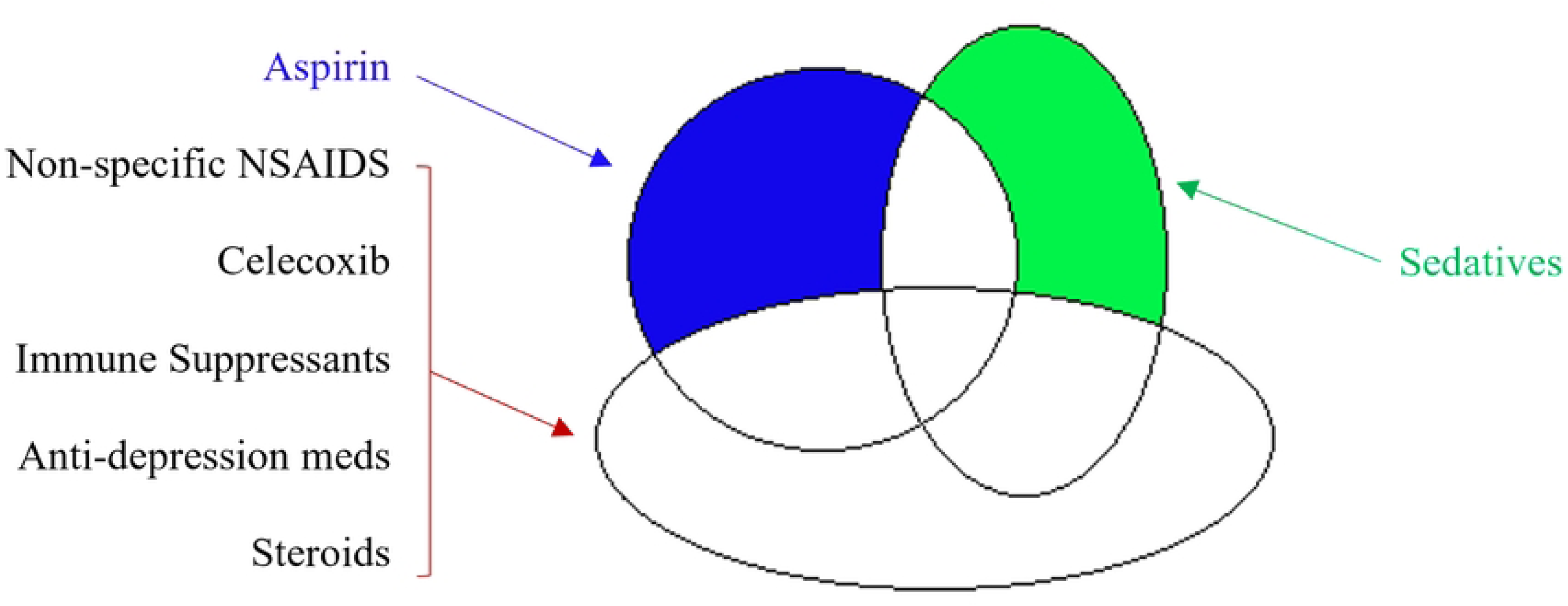
Medication-related Inclusion/Exclusion Logic for Pairwise Comparisons.

We also conducted direct outcomes comparisons for all the other pair-wise comparisons of medication groups, such as the comparison between celecoxib and aspirin. Those results appear in the S1 Supplemental Material. Those modeling results, however, are strongly transitive. For example, the result of comparing celecoxib and aspirin can be derived with considerable accuracy from the comparisons of celecoxib to the control and aspirin to the control.

Logistic regression mixed models were used to estimate M1, M2, M3, M4, M5, and M6 for each of the medication group comparisons defined in Fig 2 (a total of 36 models). In each of those models, the medication effect was defined as a binary treatment variable (e.g., celecoxib versus the control group, aspirin versus the control group, etc.). Covariates used in the models included rurality (rural, urban), gender (female, male), race (black, white, and other), ethnicity (Hispanic, non-Hispanic), age group (35-44, 45-54, 55-64, 65-74, and 75+), and Charlson Comorbidity Index [12] score (range 0 to 31), and subjects are nested within DP, and DP was incorporated into the models as a random effect. Because age was a covariate in the model, it was adjusted from the Charlson Comorbidity Index formula for this project as follow: CCI_Index = mi+ chf + pvd + stroke + dementia + pulmonary + rheumatic + pud + livermild + dm + 2*dmcx + 2*paralysis+ 2*renal + 2*cancer + 3*liverSevere + 6*mets +6*hiv. Each single disease’s contribution to this formula was also tested with stepwise logistic regression models and have no single significant effect.

Fig 4 summarizes the eligibility logic underlying the statistical modeling. Eligibility required at least one outpatient clinic visit and at least one medication prescription over a time span that ended 30 days prior to the subject’s reference date. As shown in Fig 4, we distinguished between (a) background medication use within 30 days prior to a subject’s reference date and (b) background medication use earlier than 30 days prior to the reference date. For analysis purposes, background medication group (e.g., aspirin users) included only those with evidence of background medication usage in both of those time spans (e.g., aspirin prescriptions in both time spans.)

**Figure 4:**
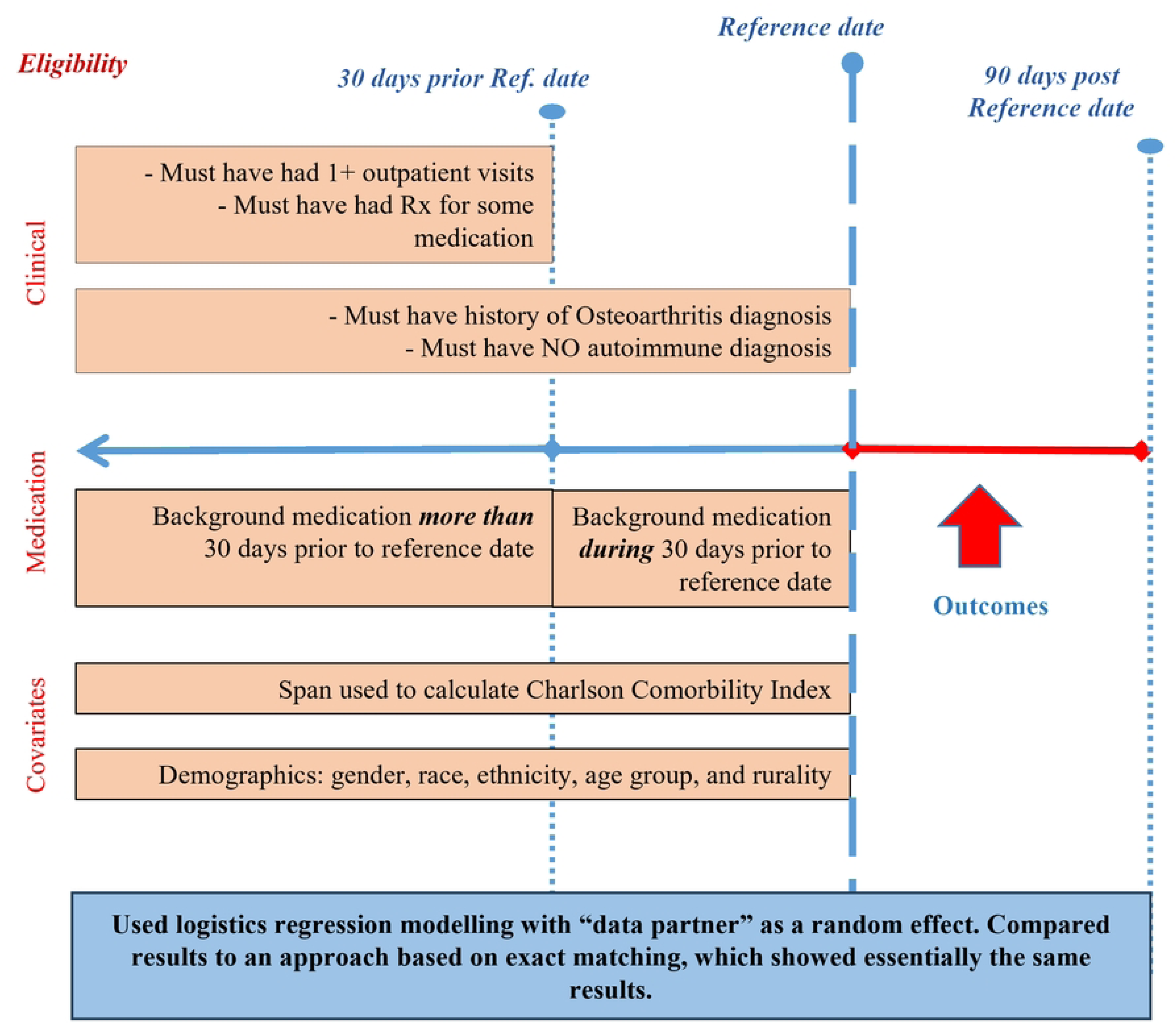
Medication Comparisons Analysis Logic.

### 2.4. Secondary Analyses

Our secondary analyses are included to help clarify interpretation of the primary analysis results. In particular, the secondary analyses attempt to address, at least to some extent, two interpretation questions:

**Question 1:** Do background medication effects estimated using data from only <u>diagnosed</u> COVID cases logically reflect the true effects of the medications?

**Question 2**: Do the primary analyses medication effects, estimated as comparisons to the control medication group, reflect effects actually generated by the background medications; or do those estimated effects simply reflect unknown risk differences that where among the factors originally determining background medication choices?

As described below, Question #1 and #2 above, both reflecting interpretation ambiguities, arise for two main reasons: (1) the fact that the background medication user groups were formed, by definition, prior to exposure to COVID, and (2) the possibility that the medication user groups differ systematically on unknown risk factors.

#### 2.4.1. Assessing for Possible Confounding due to Pre-existing Treatment Groups

The medication groups compared were implicitly formed prior to the subjects’ development of COVID. Therefore, it is plausible that the medications may have differentially affected the severity of COVID illness prior to diagnosis, leading to possible differences in the probability that an active COVID case was diagnosed. As described below, the result can be interpretation ambiguity.

Here we refer to a target medication of interest (e.g., aspirin) as the “Medication A” cohort, and the control medication users as the “Medication B” cohort. Fig 5 conceptually depicts the nested Medication A patient groups that are involved (either implicitly or explicitly) in our primary analyses. The full A1 ellipse represents the full cohort of background medication A users who meet the Fig 4 eligibility criteria, including those whose data are not on the N3C enclave, because they were never diagnosed. The A2 ellipse represents those who contracted COVID. A3 represents those who were diagnosed with COVID-19. A4 represents those who had hospital admissions due to COVID-19. And, finally, A5 represents those Medication A users who died within 90 days of COVID diagnosis. Note that, while not shown in the figure, there are analogous groups (B1, B2, B3, B4, and B5) for Medication B users (the control group.) The COVID-positive cases represented on the N3C enclave for Medication A include only those in A3 (which also implicitly includes A4 and A5). Among the Medication B cohort, the N3C enclave includes only those in B3 (which includes B4 and B5.)

**Figure 5:**
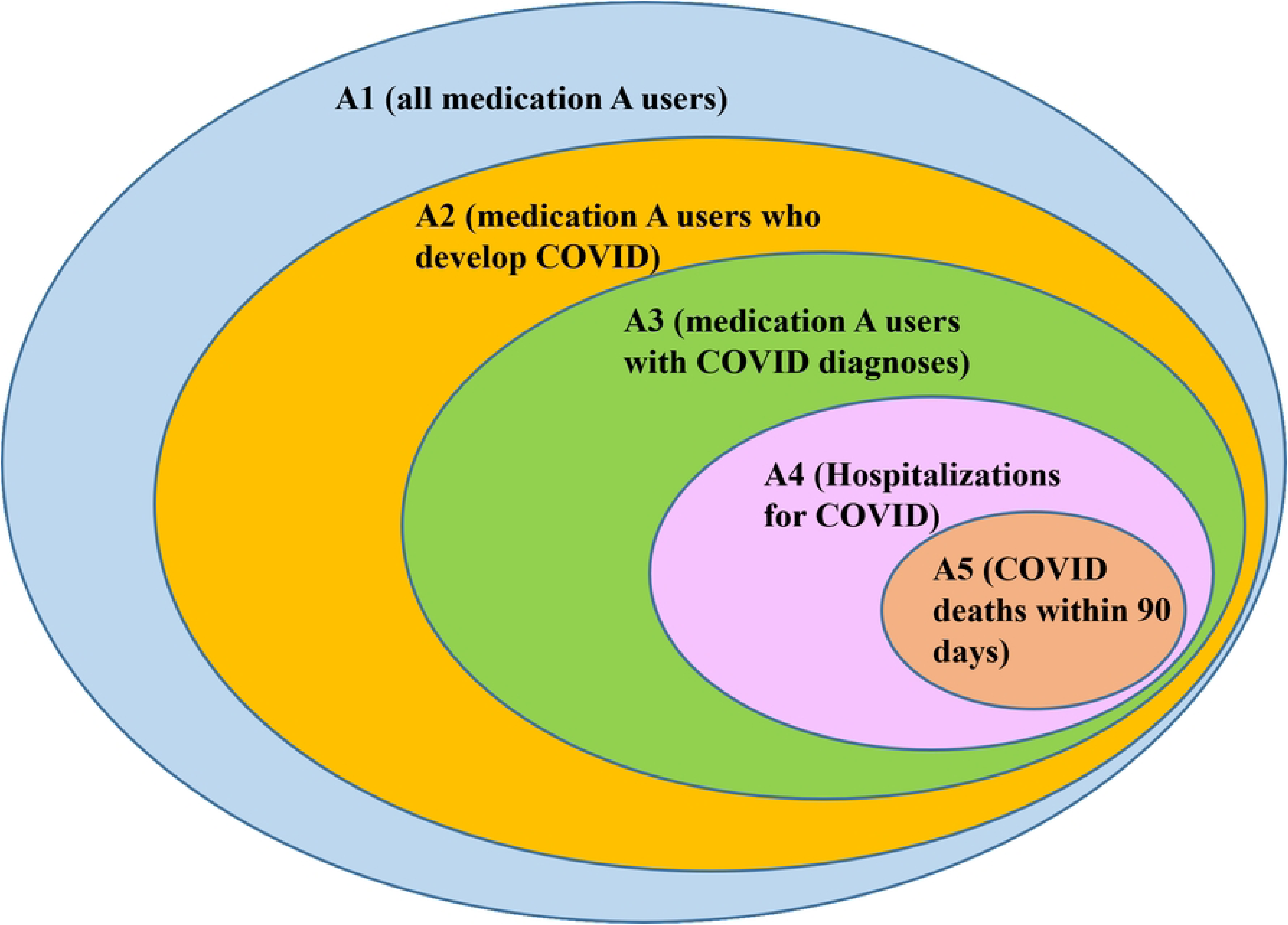
Conceptual Diagram of Nested Groups within the COVID Positive Treatment Medication Cohort.

Our primary analysis measure M1 compares the Medication A and B groups on mortality among diagnosed COVID cases. The relative risk (RR) version of M1 in terms of the Fig 5 the nested patient groups is:

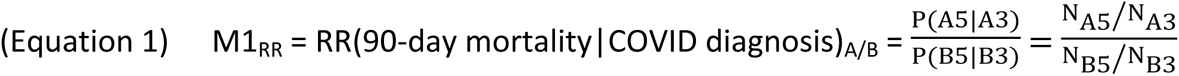

(Note: Relative Risk measures are used here, because the mathematical rationale for our secondary analyses is more efficiently described in relative risk terms.)

M1_RR_ (and M1) ignore any possible medication group difference in the probability of being diagnosed with COVID. In terms of Fig 5, M1_RR_ (and M1) use only the cases falling within the A3 and B3 sets. By contrast, a measure M7_RR_ comparing medication cohorts on full-cohort mortality would compare values of P(A5|A1) and P(B5|B1); that is:

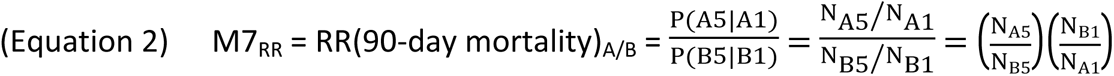

M7_RR_ also has interpretation problems; for example, it could be distorted by medication group differences in the probability of contracting COVID. However, **if it could be determined that M1_RR_ ≈ M7_RR_ holds, that would strongly suggest (albeit not conclusively prove) that pre-diagnosis medication effects are <u>not</u> distorting the medication comparison** and, therefore, M1_RR_ (and its adjusted odds ratio form M1) is a valid comparison of medication effects on COVID mortality, and is unlikely to be meaningfully distorted by medication differences in the probability of COVID diagnosis.

But measure M7_RR_ cannot be estimated directly, because NB1 and NA1 cannot be directly calculated from COVID-positive N3C data. However, estimating M7_RR_ does not necessarily require an ability to estimate N_A1_ and N_B1_ individually. All that is required is the ability to calculate the ratio N_A1_/N_B1_. The Supplemental Material describes an approach to estimating the ratio N_B1_/N_A1_ using the COVID-negative cases appearing on the N3C enclave. The results for measure M7_RR_ appearing in Section 3.0 below were obtained using that method.

Note that the above describes two different mortality constructs. M1_RR_ is a comparison of mortality among those diagnosed with COVID, while M7_RR_ is a comparison of mortality among all members of a medication usage cohort. If M1_RR_ ≈ M7_RR_ holds, the two mortality constructs are numerically very similar, in which case M1_RR_ can be viewed as a good surrogate for M7_RR_. If M1_RR_ and M7_RR_ differ substantially, it might be thought that they still are both valid measures, each being a measure for its own mortality constructs. That is true. However, as described in the Supplementary Material, it is not clear that M1_RR_ has any substantive meaning other than possibly qualifying as a surrogate for M7_RR_. The reason is that the value of M1_RR_ alone (or M1 alone) does not of itself carry any implications regarding background medication choice. M1_RR_ (and M1) may show a post-diagnosis mortality advantage for background medication A. But pre-diagnosis medication effects may negate or reverse that advantage.

Note that once M7_RR_ is estimated as described above, the measure M8_RR,_ the ratio of M7_RR_ to M1_RR_, compares the two medication groups on the probability of being diagnosed with COVID:

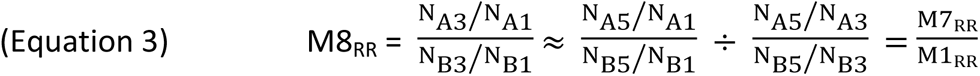

This measure also is useful in helping to clarify interpretation.

#### 2.4.2. Possible Distortion due to Missing Covariates

Each patient in a particular background medication group (e.g., the Medication A users) is presumably in the medication group for primarily clinical reasons, although patient preference, insurance coverage, and chance are contributing factors as well. It is, therefore, quite plausible that the available data are not sufficient to fully adjust for all the clinical factors underlying medication decisions.

However, in discussing the effects of missing covariates, it is helpful to first describe another potential COVID-related data characteristic; namely, that a medication’s effect on COVID outcomes may be either “indirect” or “direct.” An indirect (non-COVID-specific) effect is present if the target medication alters (reduces or increases) the risk of non-COVID outcomes by mitigating (or exacerbating) general health risk. By contrast, a direct effect on COVID outcomes is present if the target medication alters COVID outcomes risk without altering general health risk.

As described in the Supplemental Material, the medication treatment effect in a primary outcomes model (for example, the M1 effect) is a composite effect that conceptually includes:

a. Any effects of unknown risk factors that are orthogonal to the known risk factors included in the model,
b. Any indirect effects of medication A, stemming from medication A affecting the levels of risk factors, including both known and unknown risk factors, and
c. Direct effects of medication A on COVID outcomes.

The Supplemental Material describes two scenarios under which the “direct effects” can be estimated:

Scenario 1: Estimate direct effects assuming there may be unknown risk factors, but there are no indirect medication effects.

Scenario 2: Estimate direct effects assuming there may be indirect medication effects but no unknown risk factors.

As shown in Section 3.2 below (and described in the Supplemental Material), the estimated direct effect varies greatly depending on which of the above scenarios is assumed to hold, and estimates based on the above scenarios serve to conceptually bound the influence of potential unknown covariates and indirect medication effects. However, it must be kept in mind that the estimation of a medication’s direct effect is not fully satisfactory, because a medication’s indirect effect also is a true medication effect.

### 2.5 Ethics Statement

N3C operates under the authority of the National Institutes of Health IRB, with Johns Hopkins University serving as the central IRB (IRB00249128). The N3C Enclave contains de-identified, retrospective data collected by data partners (DP) under a single IRB-approved protocol through IRB reliance agreements. No data can be downloaded from the N3C enclave. Analytical code is uploaded onto the enclave, and only results can be downloaded. The study described herein was also approved by the Pennington Biomedical Research Center’s IRB (20211-016-PBRC). The N3C Data Access Committee (RP-504BA5) approved the study “COVID-19 Treatments Associated with Lower Mortality”. No informed consent was obtained from individual patients because the study used a limited data set already stripped of direct identifiers in compliance with the HIPAA Privacy Rule.

## 3. Results

### 3.1 Primary Analysis Results

Fig 6 and Table 2 summarize the results from the primary analysis models. Non-specific NSAIDs as a group were associated with significantly lower risks of admission in both the COVID-positive and the COVID-negative cohorts and with significantly lower risk of inpatient death for the COVID-negative but not the COVID-positive cohort. (For COVID-positive hospitalization rate aOR = 0.79, and for COVID-negative hospitalization rate aOR = 0.78)).

**Figure 6:**
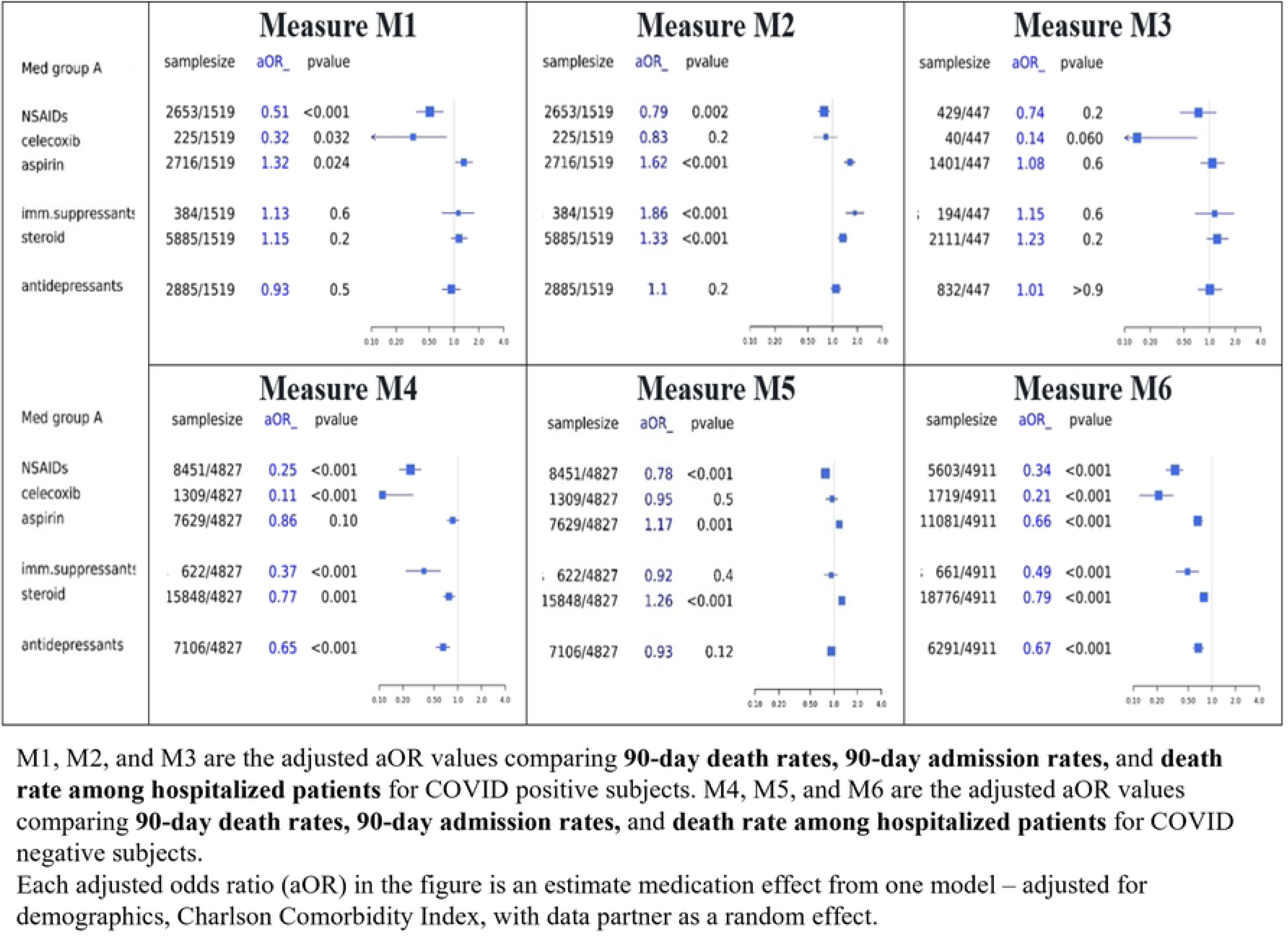
Estimate treatment effects (adjusted odds ratios) from the primary analysis.

**Table 2:** Adjusted odds ratios from the primary analysis.

| Measure | Med group A | aOR | 95% CI | p_value |
| --- | --- | --- | --- | --- |
| **M1** | NSAID | 0.51 | 0.35, 0.74 | <0.001 |
|  | Celecoxib | 0.32 | 0.09, 0.81 | 0.032 |
|  | Aspirin | 1.32 | 1.04, 1.68 | 0.024 |
|  | Immune suppressant | 1.13 | 0.72, 1.75 | 0.6 |
|  | Steroid | 1.15 | 0.93, 1.45 | 0.2 |
|  | Anti-depressant | 0.93 | 0.72, 1.19 | 0.5 |
| **M2** | NSAID | 0.79 | 0.67, 0.92 | 0.002 |
|  | Celecoxib | 0.83 | 0.59, 1.14 | 0.2 |
|  | Aspirin | 1.62 | 1.40, 1.88 | <0.001 |
|  | Immune suppressant | 1.86 | 1.45, 2.40 | <0.001 |
|  | Steroid | 1.33 | 1.17, 1.50 | <0.001 |
|  | Anti-depressant | 1.1 | 0.96, 1.26 | 0.2 |
| **M3** | NSAID | 0.74 | 0.45, 1.20 | 0.2 |
|  | Celecoxib | 0.14 | 0.01, 0.71 | 0.06 |
|  | Aspirin | 1.08 | 0.79, 1.49 | 0.6 |
|  | Immune suppressant | 1.15 | 0.68, 1.94 | 0.6 |
|  | Steroid | 1.23 | 0.93, 1.66 | 0.2 |
|  | Anti-depressant | 1.01 | 0.73, 1.40 | >0.9 |
| **M4** | NSAID | 0.25 | 0.18, 0.34 | <0.001 |
|  | Celecoxib | 0.11 | 0.04, 0.27 | <0.001 |
|  | Aspirin | 0.86 | 0.73, 1.03 | 0.1 |
|  | Immune suppressant | 0.37 | 0.22, 0.60 | <0.001 |
|  | Steroid | 0.77 | 0.66, 0.91 | 0.001 |
|  | Anti-depressant | 0.65 | 0.53, 0.79 | <0.001 |
| **M5** | NSAID | 0.78 | 0.71, 0.86 | <0.001 |
|  | Celecoxib | 0.95 | 0.80, 1.11 | 0.5 |
|  | Aspirin | 1.17 | 1.06, 1.28 | 0.001 |
|  | Immune suppressant | 0.92 | 0.75, 1.12 | 0.4 |
|  | Steroid | 1.26 | 1.16, 1.36 | <0.001 |
|  | Anti-depressant | 0.93 | 0.85, 1.02 | 0.12 |
| **M6** | NSAID | 0.34 | 0.26, 0.43 | <0.001 |
|  | Celecoxib | 0.21 | 0.12, 0.32 | <0.001 |
|  | Aspirin | 0.66 | 0.58, 0.76 | <0.001 |
|  | Immune suppressant | 0.49 | 0.35, 0.68 | <0.001 |
|  | Steroid | 0.79 | 0.70, 0.89 | <0.001 |
|  | Anti-depressant | 0.67 | 0.58, 0.78 | <0.001 |

Celecoxib showed no significant association for hospitalization in either for COVID-positive hospitalization rate aOR = 0.83, or for COVID-negative hospitalization rate aOR = 0.95. Conversely, aspirin was associated with significantly higher aOR of hospitalization in COVID-positive aOR = 1.62 and to a lesser extent in COVID-negative patients aOR = 1.17. Immune suppressants were associated with significantly higher aOR of hospitalization in COVID-positive aOR = 1.86 but not in COVID-negative cases aOR = 0.92.

Background steroid treatment was associated with significantly higher aOR of hospitalization in both groups, with very similar point estimates. For COVID-positive hospitalization rate, aOR = 1.33, and for COVID-negative hospitalization rate aOR = 1.26.

No significant associations were observed for antidepressants as a group. (For COVID-positive hospitalization rate aOR = 1.1, and for COVID-negative hospitalization rate aOR = 0.94). The medication group that showed the largest difference between COVID-positive and COVID-negative cases was immune suppressants (1.86 versus 0.92, respectively), consistent with the notion that patients on these medications are at higher risk for infectious complications, including COVID-19 [12].

When we examined the risk of death in all comers (inpatients and outpatients), non-specific NSAIDs and celecoxib were associated with significantly lower aOR of mortality in both COVID-positive (for NSAIDs’ mortality rate aOR = 0.51, and for celecoxib’s mortality rate aOR = 0.32) and COVID-negative cases (for NSAIDs’ mortality rate aOR = 0.25, and for celecoxib’s mortality rate aOR = 0.11), while aspirin was associated with significantly higher aOR of mortality in COVID-positive aOR = 1.32 but not in COVID-negative aOR = 0.86. Similarly, background steroid treatment was associated with significantly higher aOR of mortality in COVID-positive aOR = 1.15 but not n COVID-negative cases aOR = 0.77 while antidepressants showed no significant association with aOR of mortality in COVID-positive aOR = 0.93 and a significant negative association in COVID-negative aOR= 0.65.

### 3.2 Secondary Analysis Results

The secondary analyses focus on the interpretation questions posed in Section 2.4. Fig 7 compares values of measure M1_RR_ (relative risk of death given a COVID diagnosis) to values of M7_RR_ (relative risk of COVID death among all medication cohort members.) Results for the two measures are quite similar for all the medications except celecoxib and the immune suppressants, for which M1_RR_ and M7_RR_ differ approximately by a factor of two, but in different directions. The implication is that the diagnosis-denominated measure, M1_RR_, may not adequately reflect the COVID mortality experience of those two medication groups. However, the sample sizes for celecoxib and for immune suppressants are too small to draw any confident conclusions.

**Figure 7:**
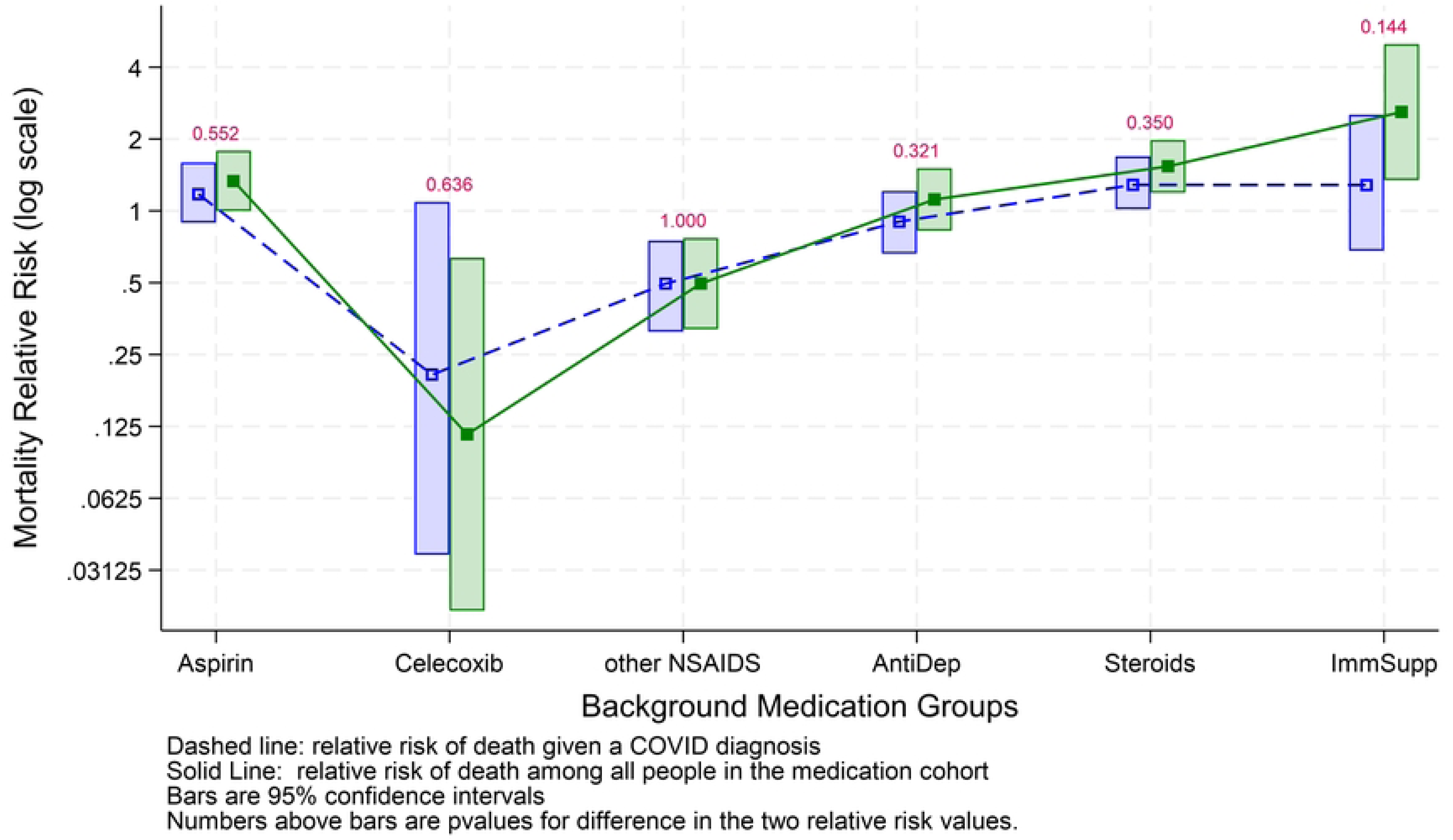
Relative Risk of Death|COVID Diagnosis and Relative Risk of Death|in Medication Cohort.

Fig 8 shows estimates of medication direct effects under two different assumptions: an assumption that some relevant covariates are unknown but there are no indirect medication effects, versus the assumption that indirect medication effects may be present but there are not missing risk factors. The adjusted odds ratios for our primary analysis measure M1 are graphed by the solid black line. The dashed red line graphs M1 after adjustment for the potential effects of unknown risk factors. The dashed blue line graphs M1 after adjustment for the potential indirect medication effects. Of course, bias may be present from both unknown covariates and indirect medication effects. The potential bias from indirect medication effects is much less than the potential bias from unknown covariates. Further, the <u>actual</u> bias present from either source may be much less than the full <u>potential</u> bias quantified in Fig 8.

**Figure 8:**
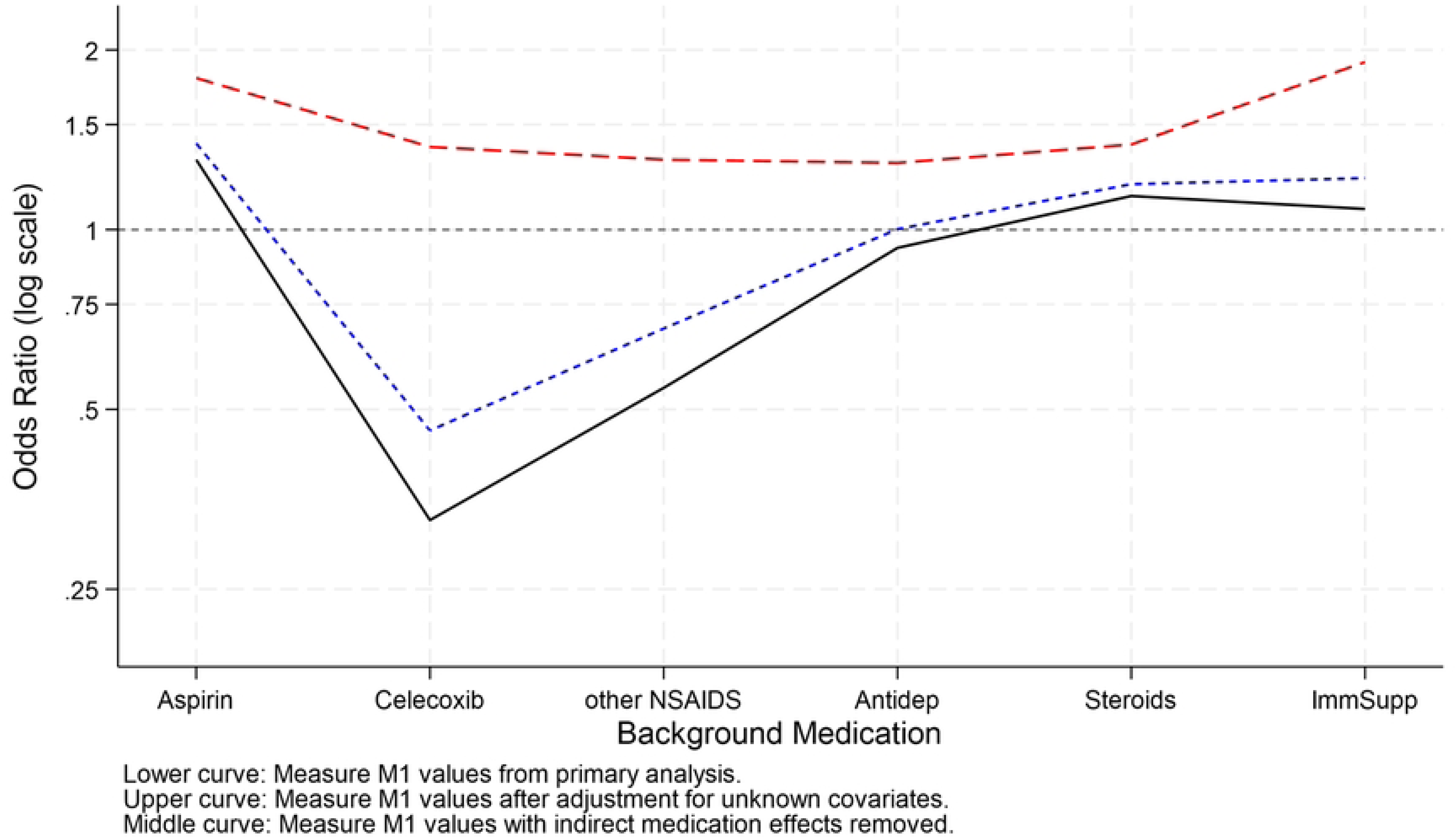
Medication Effects after adjustment for unknown covariates of adjustment for indirect effects.

In Fig 8, results under an assumption of indirect medication effects seem more plausible than results assuming possible missing covariates.

## 4. Discussion

With the interpretation caveats described above, given osteoarthritis patients who had not been diagnosed with autoimmune disorders as a target population, background NSAIDs except aspirin were associated with lower hospitalization rates as compared to the non-anti-inflammatory control group (sedatives). In contrast, background aspirin and systemic steroids were associated with higher admission rates compared to control.

The results described in this study in the N3C enclave provide potentially useful information for clinicians treating patients at risk for COVID-19. It is important to point out that these results do not necessarily imply that pharmacological activities of the agents we studied directly affect COVID-19 outcomes. This is because of the interpretation challenges described under Methods, and because groups of patients identified by treatment with each of these medication classes do not have identical baseline risks of hospitalization and death, nor do they have identical baseline risk profiles for SARS-CoV2 infection or symptomatic COVID-19 disease. Rather, each medication class can be seen as identifying a group of patients potentially at higher or lower risk of hospitalization or death, and COVID-19 adds to these different baseline risk profiles. Among patients with osteoarthritis, background prescription NSAIDs, either non-specific or COX-2 selective, do not appear to contribute to the risk of COVID-19 hospitalization or death, with the prominent exception of aspirin. Patients on background prescription aspirin appear to be at higher risk of hospitalization when COVID-positive, and to a lesser extent when COVID-negative, and to be at significantly higher risk of non-inpatient death (presumably from acute complications prior to hospitalization) when COVID-positive but not when COVID-negative. A possible explanation for this is protopathic effect, meaning that aspirin is used to treat conditions that increase the risk of hospitalization and death in COVID-positive patients. Unlike other NSAIDs, in addition to its uses as an anti-inflammatory agent, aspirin is used alone or in combination with other agents as part of anti-platelet therapy to prevent thrombosis in patients at risk after acute cardiovascular events or procedures, e.g. patients with percutaneous coronary intervention (PCI), [13], total hip arthroplasty [14], or limb revascularization [15]. Furthermore, prescription aspirin is used for the primary prevention of coronary artery disease [16]. These conditions are likely to increase the baseline risk of hospitalization and sudden death in COVID-positive patients. Cardiovascular disease (CVD) is a recognized risk factor for COVID-19 morbidity and mortality [17]. Our data suggest that patients on chronic prescription aspirin who are still unvaccinated against currently circulating strains of SARS-CoV-2 should consider COVID-19 vaccination and mitigate the risk of infection.

Our results for NSAIDs are consistent with the results obtained by Reese et al [11], whose estimated effect of NSAIDs, including Celecoxib, on inpatient mortality (OR = 0.51) lies between our estimated effects for Celecoxib (OR = 0.14) and non-specific NSAIDs (OR = 0.74).

Celecoxib deserves separate consideration, as background treatment with it was associated with reduced relative risk of death in both COVID-positive and COVID-negative patients. We do not know whether these effects are due to a hidden variable common to celecoxib-treated patients or can be attributed to the medication itself. If celecoxib simply masked symptoms of mild COVID-19, preventing diagnosis of such cases, one would expect the opposite effect, i.e., a higher risk of severe outcomes in celecoxib-treated patients. COX-2 inhibition had been proposed as a possible treatment for COVID-19 [18] and a small clinical trial in mild or moderate COVID-19 supports this notion [19]. In that study, lymphocyte counts were increased in patients taking celecoxib. Furthermore, a large real-world evidence observational study by Liao [20], based on claims data from 2,935,415 unvaccinated and 189,692 vaccinated patients, identified NSAIDs including celecoxib-aspirin combinations, celecoxib-ibuprofen combinations and celecoxib single agent as having repurposing potential for COVID-19 treatment [20]. Unlike our results, aspirin was also associated with a lower risk of COVID-19 mortality in that study. Significant differences between the two studies are the inclusion criteria, with in our study are restricted to osteoarthritis without autoimmune disorders, and the fact that risk adjustment in Liao’s study used a proprietary Optum Episode Risk Group Score [18]. A different theoretical approach, using multi-evidence deep graph neural networks [21], predicted anti-COVID-19 activities for aspirin and celecoxib. A molecular modeling study identified celecoxib as a potential inhibitor of the SARS-CoV-2 main protease, thus hypothesizing a direct antiviral activity [22]. Besides this putative antiviral activity, COX-2, the primary target of celecoxib, is a key player in inflammation, and its major product, PGE_2_, suppresses T-cell-mediated cellular immunity while promoting Th2 responses [23]. It is possible that COX-2 inhibition may improve immune responses to SARS-CoV-2. The apparent association of background celecoxib with lower risk of mortality in COVID-negative patients is more difficult to explain. Given that celecoxib is contraindicated in patients with history of cardiovascular events, including myocardial infarction and PCI or other coronary revascularization, it is possible that the baseline risk of cardiovascular events in patients on celecoxib may be lower than that of patients on other medication groups we studied. However, the PRECISION trial results showed that at currently recommended doses, the cardiovascular and renal safety profile of celecoxib is favorable compared to non-specific NSAIDs ibuprofen and naproxen [24]. It’s worth noting that PGE_2_ also suppresses tumor immunity [25, 26], some studies support the use of celecoxib as an adjuvant to cancer treatment [27] and a vast literature supports the use of NSAIDs [28–31], and celecoxib in particular [32] as cancer chemo-preventive agents. Initial enthusiasm for selective COX-2 inhibitors was dampened by their cardiovascular side effects [33]. Our data, within the limitations of the N3C dataset and the analytical strategy we used, are consistent with the notion that background use of celecoxib is associated with a decreased risk of all-cause mortality in COVID-negative and -positive patients. This data supports the predictions made by Liao [20] and Hsieh et al. [21].

Background steroid medications were association with increased risk of inpatient death and all-comers death in COVID-19 positive but not in COVID-19 negative patients. Despite the fact that dexamethasone is effective in treating severe COVID-19, it is possible that chronic use of steroids may dampen protective immunity against SARS-CoV-2.

Immune-suppressants (e.g. rapalogs) were associated with increased risk of hospitalization but not death in COVID-19 positive but not negative cases. These medications are usually prescribed to transplant recipients, who have been identified as being at higher risk for severe COVID-19 [32]. A possible explanation for our findings is that patients on immune-suppressants on average adopted precautions to avoid SARS-CoV-2 exposure, but once infected were more likely to be hospitalized. The lack of significantly increased risk of death is an encouraging finding.

## 5. Conclusions

In conclusion, our analysis of N3C enclave data suggests that patients on prescription non-specific NSAIDs or celecoxib do not face increased risks of severe outcomes in COVID-19, with the possible exception of patients on aspirin. Celecoxib was associated with decreased risk of mortality in both COVID-negative and COVID-positive cases. Unvaccinated patients on chronic systemic steroids and particularly immune-suppressive medications may face increased risk of severe COVID-19 outcomes. Our data do not support the notion that background treatment with antidepressants or fluvoxamine affect COVID-19 outcomes in the patient categories we studied.

## Data Availability

Data cannot be shared because The N3C Data Enclave is managed under the authority of the NIH information can be found at https://ncats.nih.gov/n3c/resources.

## Acknowledgements

National COVID Cohort Collaborative (N3C) Consortium membership includes:

1. Christopher G. Chute

Johns Hopkins University

## Contributions

clinical data model expertise, data curation, data integration, data quality assurance, data security, funding acquisition, governance, N3C Phenotype definition, project evaluation, project management, and regulatory oversight/admin.

## Individual Acknowledgements for Core Contributors

We gratefully acknowledge the following core contributors to N3C: Adam B. Wilcox, Adam M. Lee, Alexis Graves, Alfred (Jerrod) Anzalone, Amin Manna, Amit Saha, Amy Olex, Andrea Zhou, Andrew E. Williams, Andrew M. Southerland, Andrew T. Girvin, Anita Walden, Anjali Sharathkumar, Benjamin Amor, Benjamin Bates, Brian Hendricks, Brijesh Patel, G. Caleb Alexander, Carolyn T. Bramante, Cavin Ward-Caviness, Charisse Madlock-Brown, Christine Suver, Christopher G. Chute, Christopher Dillon, Chunlei Wu, Clare Schmitt, Cliff Takemoto, Dan Housman, Davera Gabriel, David A. Eichmann, Diego Mazzotti, Donald E. Brown, Eilis Boudreau, Elaine L. Hill, Emily Carlson Marti, Emily R. Pfaff, Evan French, Farrukh M Koraishy, Federico Mariona, Fred Prior, George Sokos, Greg Martin, Harold P. Lehmann, Heidi Spratt, Hemalkumar B. Mehta, J.W. Awori Hayanga, Jami Pincavitch, Jaylyn Clark, Jeremy Richard Harper, Jessica Yasmine Islam, Jin Ge, Joel Gagnier, Johanna J. Loomba, John B. Buse, Jomol Mathew, Joni L. Rutter, Julie A. McMurry, Justin Guinney, Justin Starren, Karen Crowley, Katie Rebecca Bradwell, Kellie M. Walters, Ken Wilkins, Kenneth R. Gersing, Kenrick Cato, Kimberly Murray, Kristin Kostka, Lavance Northington, Lee Pyles, Lesley Cottrell, Lili M. Portilla, Mariam Deacy, Mark M. Bissell, Marshall Clark, Mary Emmett, Matvey B. Palchuk, Melissa A. Haendel, Meredith Adams, Meredith Temple-O’Connor, Michael G. Kurilla, Michele Morris, Nasia Safdar, Nicole Garbarini, Noha Sharafeldin, Ofer Sadan, Patricia A. Francis, Penny Wung Burgoon, Philip R.O. Payne, Randeep Jawa, Rebecca Erwin-Cohen, Rena C. Patel, Richard A. Moffitt, Richard L. Zhu, Rishikesan Kamaleswaran, Robert Hurley, Robert T. Miller, Saiju Pyarajan, Sam G. Michael, Samuel Bozzette, Sandeep K. Mallipattu, Satyanarayana Vedula, Scott Chapman, Shawn T. O’Neil, Soko Setoguchi, Stephanie S. Hong, Steven G. Johnson, Tellen D. Bennett, Tiffany J. Callahan, Umit Topaloglu, Valery Gordon, Vignesh Subbian, Warren A. Kibbe, Wenndy Hernandez, Will Beasley, Will Cooper, William Hillegass, Xiaohan Tanner Zhang. Details of contributions available at covid.cd2h.org/core-contributors.

The authors received direct benefit from discussions and presenting the project to the N3C – Rural Health Domain teams. The authors would like to thank Sharon Patrick and the West Virginia Clinical and Translational Science Institute (WVCTSI) for project planning and management.

## Supporting information

**S1 Supplemental Material**

